# Social Listening: A Thematic Analysis of COVID-19 Discussion on Social Media

**DOI:** 10.1101/2020.07.25.20162040

**Authors:** Sulaimon Afolabi, Sakinat Oluwabukonla Folorunso, Zinia Siphosethu Bunyula, Oluwatobi Oluwaseyi Banjo, Sibusiso Sydney Matshika, Warrie Usenobong Warrie, Naledi Ngqambela, Ayodeji Emmanuel Adepoju, Hendrica Rabophala, Olawale Victor Abimbola, Michael Segun Olanipekun, Adedayo Lateef Odukoya

## Abstract

COVI-19 is a variant of coronavirus diseases that has destabilised the entire world and whose cure as at mid-2020 has become elusive. Social media is ablaze with discussions around the pandemic. There is the dire need to delineate the on-going conversations on the infection with the intention of creating awareness on people’s reaction, opinion, action and recommendation that are inimical to the wellbeing of the populace. Hence, this study is geared towards performing thematic analysis of the discussions on social media about COVID-19. We programmatically retrieved data from twitter between 1st March, 2020 to 30th June 2020 with covid-19 related keywords. We processed the data and later categorised them into themes that evolved from the tweets namely Herbs and Vegetables as COVID-10 Panacea, Self-Medication Due to Prescription by Non-Medical Practitioners on Social Media, Conspiracy Theories on COVID-19 and Fear and Anxiety Associated with COVID-19. The results show that many are resulting to herbs to protect themselves against the disease; taking drugs without doctor’s prescription; believing in conspiracy theories and having certain degree of fear.

## Introduction

COVI-19 is a variant of coronavirus diseases that has destabilised every facet of human endeavour including education, sport, business, tourism, transportation, religious and social activities since its emergence in late 2019 (Chang, Yan, & Wang, 2020; Lodigiani et al., 2020; Nicola et al., 2020; Ozili & Arun, 2020; Torales, O’Higgins, Castaldelli-Maia, & Ventriglio, 2020). The cure as at mid-2020 has become elusive and the stringent measures that different governments across the globe have adopted to curb its spread are seemingly not lasting palliative as the infection rate goes on the increase as soon as the strict rules are removed. Chief among the rules is the total locked down of some of the affected cities or countries. Since its onset in late 2019 in Wuhan, China, the novel coronavirus has spread rapidly to different part of the world and it has killed thousands of its hosts (Baud et al., 2020; Jordan, Adab, & Cheng, 2020; Weiss & Murdoch, 2020).

COVID-19, just like the other types of coronavirus e.g. severe acute respiratory syndrome (SARS) and Middle East respiratory syndrome (MERS) negatively impact the human respiratory organ. And the symptoms of the disease are not limited to the respiratory system malfunction and they are numerous. The symptoms include the following; runny nose, headache, dry cough, sore throat, fever, diarrhoea, tiredness, skin rash, loss of taste and so on (Huang & Zhao, 2020; Rothan & Byrareddy, 2020). Furthermore, it is interesting to note that not infected individuals manifest the symptoms. That is, they are asymptomatic most especially at the early stage of contracting the disease. The virus can be transmitted or contracted through direct contact with the carrier’s droplets, touching of contaminated surface including aerosolized and faecal-oral transmission.

Due to the global spread of the infectious disease, Africa has not been exempted from the virus due its openness to international trade and migration. Due to the state of its public health, it is believed that Africa lacks the capacity to detect and contain the pandemic and probably the worst affected Africa will be worst affected continent (Gilbert et al., 2020; Nkengasong & Mankoula, 2020). In Africa, the first confirmed COVID-19 case was on the 14 February 2020 in Egypt. Shortly after this case, several infections were confirmed by 31 March 2020, infections in Africa have been following an upward trend and exceeded five hundred thousand cases. The impact of the infection in Africa does not differ from what is obtainable in other continents as its social, health and economic activities has been negatively affected.

Social media is ablaze with discussions around the pandemic and these conversations are often characterised by a number of traits suggesting that those utterances are worthy of closer examination. First, fear and anxieties are common expressions on various social media outlets as the devastating impacts of coronavirus are being relayed via the same platforms (Garfin, Silver, & Holman, 2020; Kumar & Somani, 2020; Lin, 2020). Although, it is normal for people to express fear in the face of ravaging disease such as COVID-19, the disadvantage of this is that this action predisposes them to making trying all the suggestions or panacea that come their way as we shall see in the subsequent sentences. Second, there are suggested precautionary measures that are ridden with myth that are being propagated on various social media platforms which those who are perplexed by the epidemics can easily subscribe to at their own demerit. For instance, it has been suggested widely that eating certain herbs and vegetables such as black pepper, ginger, garlic, lemon and other herbal products can prevent against getting infected with COVID-19 but this has been refuted (Check, 2020; Rosenbloom, 2020; Wingard, 2020). While there is a general consensus on the health importance of the listed items, however, they do not possess enough potency to either fight nor prevent people from being susceptible to the disease. Third, certain conspiracy theory on the fact that 5g technologies specifically mast is one form of transmitting the virus to the populace has led to violent protests and reactions across the globe and this has resulted to loss of lives and properties. Various researchers in the field telecommunication field has not established any connection between 5g technologies and COVID-19 (Ahmed, Vidal-Alaball, Downing, & Seguí, 2020; Uthman et al., 2020). Fourth, extreme climatic conditions are inimical to the transmission of COVID-19. Weather condition such as extremely hot, humid or cold does not protect from contracting the disease. This statement is not true as

Most of the existing literatures studied the impact of coronavirus on Global health, Education or sport without considering the opinions of people on social media. It has been reported that people share their opinions, feelings or emotion on social media. Therefore, studying the opinions of people on social media about coronavirus is very important. The objectives of this paper are as follows

## Method

In order to achieve the purpose of this study, we adopted thematic analysis, which is a type of qualitative method of study. Also, we sourced the data for this study from twitter between 1st of March and 30th of June. Our data is in English and not limited to any geographical location. The text data extraction was done in a systematic way as we retrieved tweets by theme. The following are the themes that we extracted the data based on. (1) Herbs and vegetables as COVID-19 panacea theme data was extracted using keywords such as garlic, ginger, tumeric, lemon, pepper, honey in combination with either coronavirus or covid e.g. ginger and covid. All these individual datasets were later combined and duplicate records were removed (ii) COVID-19 Self-medication based ono prescription by non-medical practitioners on social media theme data was extracted similarly to the first theme with the following key words hydroxychloroquine, plaqueil, chloroquine, coronavirus, self-medication, antibiotics, coronavirus and covid (iii) Conspiracy theories on COVID-19 theme data was extracted using the following criteria 5G, conspiracy, china lab, Bill Gates in combination with COVID and coronavirus and duplicate records were removed. (iv) Fear and anxiety mongering theme data was downloaded for the same study period using fear, afraid, panic, feeling, covid, anxious, anxiety in combination with either covid and coronavirus. We performed thematic analyses on the tweets by further creating sub-themes from the major ones. The themes and subthemes are shown in Table 1.

**Table 1:**
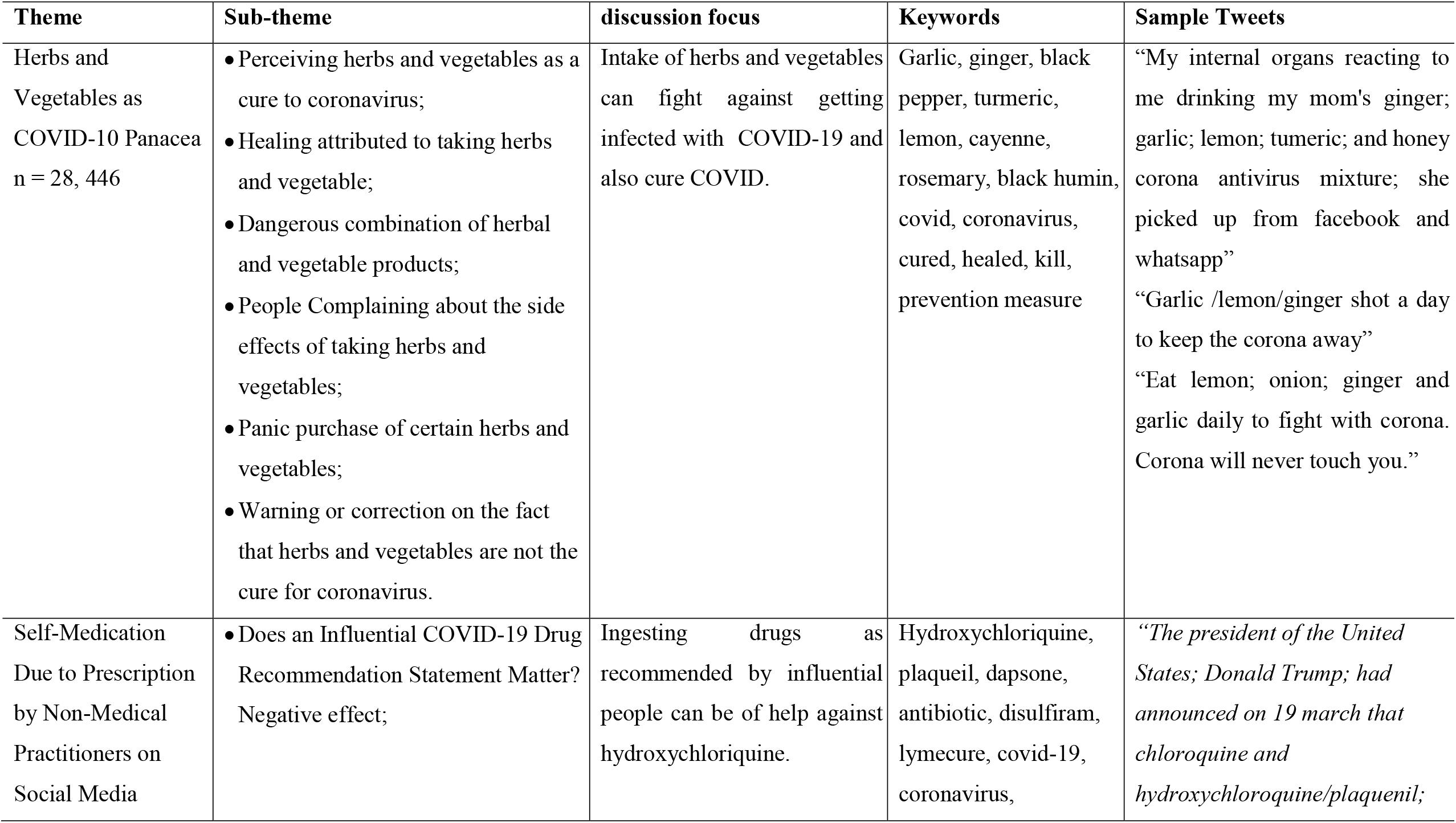
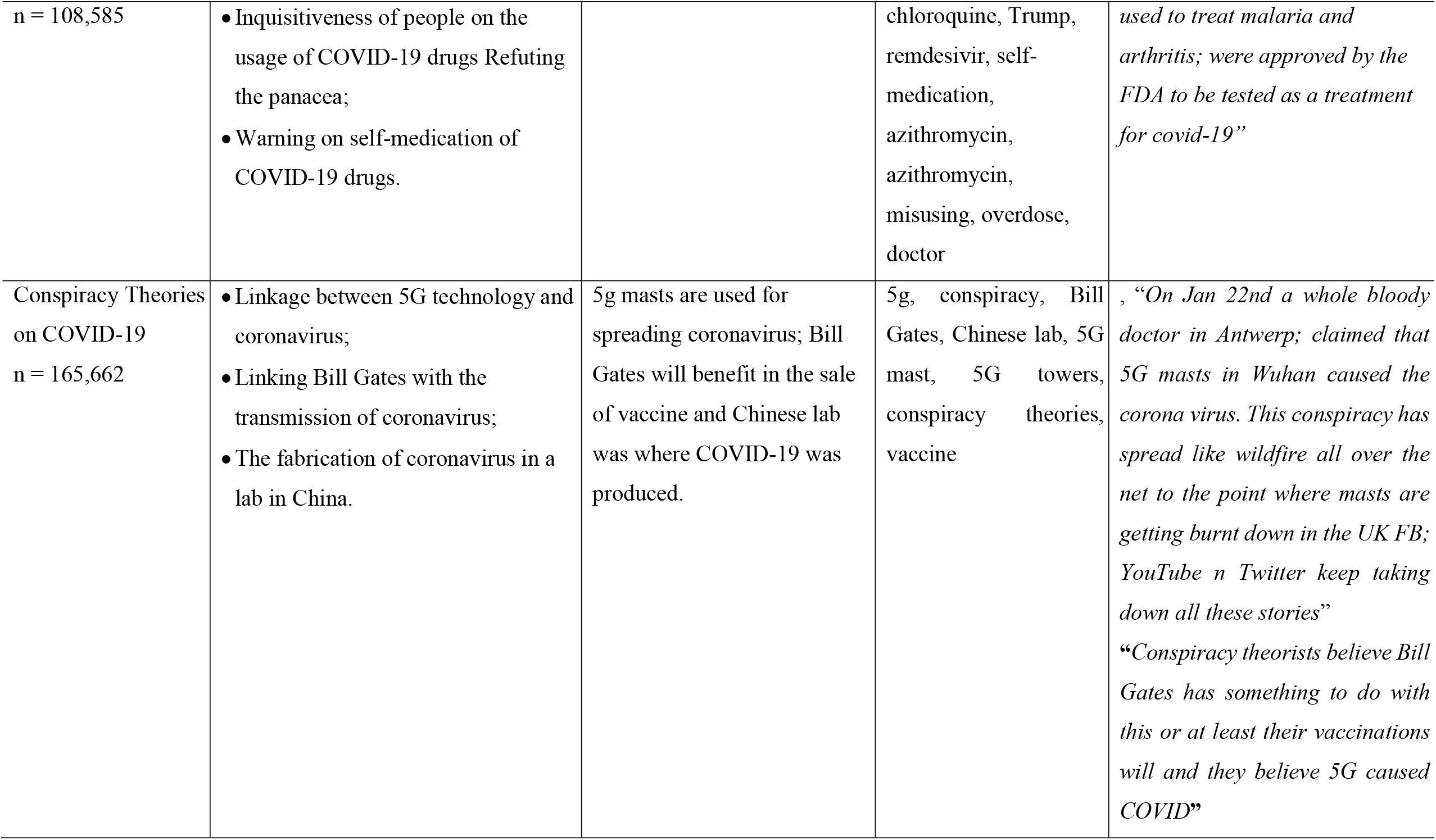
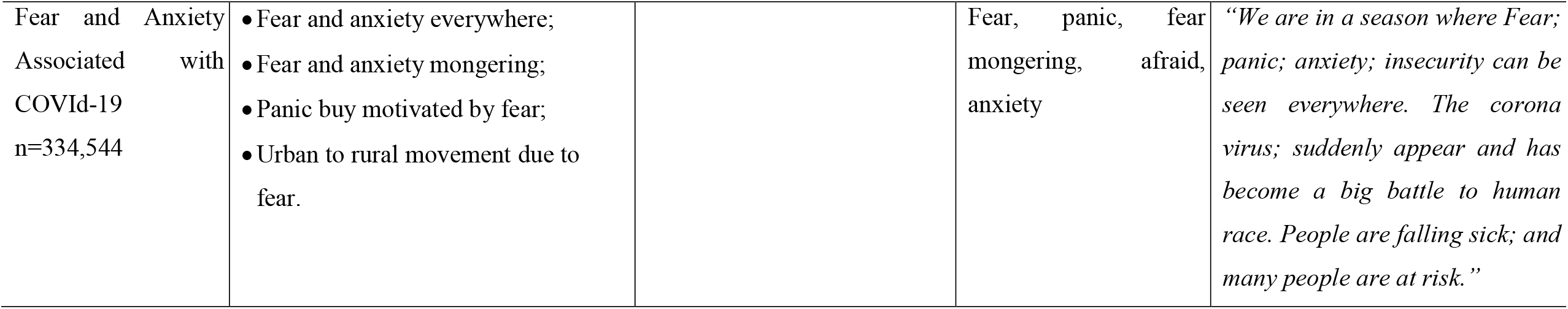
Themes, sub-themes, discussion focus, keywords and sample tweets on twitter discussion around the pandemic

| Theme | Sub-theme | discussion focus | Keywords | Sample Tweets |
| --- | --- | --- | --- | --- |
| Herbs and Vegetables as COVID-10 Panacea<br>n = 28, 446 | • Perceiving herbs and vegetables as a cure to coronavirus;<br>• Healing attributed to taking herbs and vegetable;<br>• Dangerous combination of herbal and vegetable products;<br>• People Complaining about the side effects of taking herbs and vegetables;<br>• Panic purchase of certain herbs and vegetables;<br>• Warning or correction on the fact that herbs and vegetables are not the cure for coronavirus. | Intake of herbs and vegetables can fight against getting infected with COVID-19 and also cure COVID. | Garlic, ginger, black pepper, turmeric, lemon, cayenne, rosemary, black humin, covid, coronavirus, cured, healed, kill, prevention measure | “My internal organs reacting to me drinking my mom's ginger; garlic; lemon; tumeric; and honey corona antivirus mixture; she picked up from facebook and whatsapp”<br>“Garlic /lemon/ginger shot a day to keep the corona away”<br>“Eat lemon; onion; ginger and garlic daily to fight with corona. Corona will never touch you.” |
| Self-Medication Due to Prescription by Non-Medical Practitioners on Social Media | • Does an Influential COVID-19 Drug Recommendation Statement Matter? Negative effect; | Ingesting drugs as recommended by influential people can be of help against hydroxychloriquine. | Hydroxychloriquine, plaqueil, dapsone, antibiotic, disulfiram, lymecure, covid-19, coronavirus, | *“The president of the United States; Donald Trump; had announced on 19 march that chloroquine and hydroxychloroquine/plaquenil;* |
| n = 108,585 | • Inquisitiveness of people on the usage of COVID-19 drugs Refuting the panacea;<br>• Warning on self-medication of COVID-19 drugs. |  | chloroquine, Trump, remdesivir, self-medication, azithromycin, azithromycin, misusing, overdose, doctor | *used to treat malaria and arthritis; were approved by the FDA to be tested as a treatment for covid-19”* |
| Conspiracy Theories on COVID-19<br>n = 165,662 | • Linkage between 5G technology and coronavirus;<br>• Linking Bill Gates with the transmission of coronavirus;<br>• The fabrication of coronavirus in a lab in China. | 5g masts are used for spreading coronavirus; Bill Gates will benefit in the sale of vaccine and Chinese lab was where COVID-19 was produced. | 5g, conspiracy, Bill Gates, Chinese lab, 5G mast, 5G towers, conspiracy theories, vaccine | *, “On Jan 22nd a whole bloody doctor in Antwerp; claimed that 5G masts in Wuhan caused the corona virus. This conspiracy has spread like wildfire all over the net to the point where masts are getting burnt down in the UK FB; YouTube n Twitter keep taking down all these stories”*<br>*“Conspiracy theorists believe Bill Gates has something to do with this or at least their vaccinations will and they believe 5G caused COVID”* |
| Fear and Anxiety Associated with COVID-19<br>n=334,544 | • Fear and anxiety everywhere;<br>• Fear and anxiety mongering;<br>• Panic buy motivated by fear;<br>• Urban to rural movement due to fear. |  | Fear, panic, fear mongering, afraid, anxiety | *“We are in a season where Fear; panic; anxiety; insecurity can be seen everywhere. The corona virus; suddenly appear and has become a big battle to human race. People are falling sick; and many people are at risk.”* |

## Results

We searched for tweets of interest using certain criteria between 1st of March to June 30 2020. For herbs and vegetables as COVID-19 theme, twitter text data extraction produced 28,446 unique records with the following key words ginger, garlic, tumeric, lemon, pepper, honey, coronavirus and COVID used as the search criteria. We extracted 108, 586 for theme self-medication due to prescription by non-medical practitioners on social media using the following keywords hydroxychloroquine, plaqueil, chloroquine, coronavirus, self-medication, antibiotics, coronavirus and COVID. For the theme on COVID-19 related conspiracy theories we extracted 174,575 records based on the following criteria 5G, conspiracy, china lab, Bill Gates, COVID and coronavirus. We extracted a total of 334,544 records for Fear and Anxiety Associated with COVId-19 with the key words such fear, panic, fear mongering, afraid, anxiety. Table 1 provides summary information on the information statement above including the following themes, subthemes, myths, keywords and sample tweets.

### Herbs and Vegetables as COVID-19 Theme

This theme fused many diverse range of sentiments on using herbs and vegetables to prevent being infected with COVID-19 virus getting cured with the concocted herbs and vegetables when sick; getting false sense of cure when people are no longer feeling the symptoms. Some people even take dangerous combination of these herbs which could have side effects in the long run with a section warning of the futility of taking the concoction as it neither shields or cure people of the infection.

Sentiments of people around taking herbs and vegetables as COVID-19 as a prevention: There are people who believed that taking herbs and vegetable can actual protect against getting infected with the disease. To them, it is a form vaccine. You will find making statements like, “Eat lemon; onion; ginger and garlic daily to fight with corona. Corona will never touch you”; *“Corona virus, slices of lemon in a cup of hot water can save (yo)ur lift”; “Because we are invincible with haldi, honey, neem, tulsi etc. and corona cannot touch us*.*”; “I am craving for hot pepper soup to scare bro corona”; “At a bar yesterday; someone said; hot spicy pepper soup is a preventive measure/cure for the corona virus”; “Preventive measure for corona: drink hot water with citric acid or lemon juice”. “Increase your immunity by boiling garlic with ginger; add lemon and honey drink a cup twice or thrice daily. no cough; no respiratory infections; even corona will be rendered impotent. a simple therapy that works. you can add tumeric as one of the ingredient”; “i have been taking it on and off for years; month on; month off but since covid-19 has been let loose I’ve upped my dosage along with paracetamol. I’m not saying it’s a cure but short of vaccination it’s about the best you can do*…*along with ginger; garlic”* From all indications a number of people on the social media platform are advocating for the intake of farm produce such garlic, ginger, pepper, honey and so to fight against COVID-19.

#### Perceiving herbs and vegetables as a cure to coronavirus

Many people on twitter believed that in actual fact that vegetables and herbs are the solution to the problem of coronavirus and some seem to be sure about this. They even encourage other to share the solution. *“organic tumeric; ginger; honey; garlic and lemon will cure COVID 19. Spread the word”*. It is interesting to note that some provide the full procedure to make the herbal solution to be potent. “*Cure for COVID -19, lemon; garlic; ginger; and tumeric powder. Boil the mixture for 10-15 mins. Pour it into a bucket; place your head slightly above the bucket and cover your head with a blanket so that the heat does not escape. inhale the vapour for about”; “God has provided natural cure for every disease. we need to go natural. garlic; tumeric and ginger can handle covid-19. Use it as spices in every meal. make chicken pepper soup with it. Chew it 3 times daily. make fresh juice with it*.*”*

#### Healing attributed to taking herbs and vegetables

There are people with testimonies expressing that they become healed after taking herbs and vegetables but this cannot be verified “I *went through COVID had all the symptoms; then recovered after 2 weeks; made sure I had black seed oil with ginger; garlic; lemon and honey*.”; “*we healed two of housemates of covid-19 with thyme tea; probiotics; black seed oil; garlic; ginger; fresh juices; rest; seamoss; bladder wack; water and patience*.”. Some of the testimonies originated from influential people as they occupy high position in the government of their respective country. For instance, “*Tanzania president just said ginger and lemon healed his son from corona so what’s y’all ghat to say about that*…*”; “Tanzanian president Magufuli revealed that his own child had contracted the [covid-19] virus but was now well and ‘doing push-ups*.*’ he said the child had made a recovery following a regimen of self-isolation; steam inhalation; and lemon and ginger”; “Seyi Makinde (a governor in Nigeria) revealed that he took carrots and honey then he was healed of corona virus whereas people are dying in US. the whole thing is a scam”; “He took black seed oil, honey, carrot and vitamin C to fight corona virus, Oyo state governor, Seyi Makinde”*. Some have confessed to the fact they have not only one but multiple people being healed. “*My mum has sent me at least 5 testimonies from supposed COVID-19 ‘survivors’ that all swear ginger + garlic + honey + lemon is what cured them*”; “*I don’t know how true this is but trying it won’t be bad. This simple remedy; has cured over 5 corona virus patients in the UK last week. slice small garlic; ginger; onion; salt; lemon; add small water*”

#### Dangerous combination of herbal and vegetable products

We heard people saying that they combined many of these items with possibility of food poisoning. Read these statements, “*Corona fayras made biggest blunder of his life by visiting Somalia. Wait & see how deadly combinations of xabad sowda (black cumin seed oil),sanjibil (ginger),filfil (black pepper),toon(garlic) & tahlil will send it back to Wuhan*”; “*Pls tell her to start treating herself with local remedy*.. *ginger; garlic; turmeric and lemon blend together*… *This combination contains a lot of zinc and vitamins to improve her immune system to fight any virus including COVID 19*…*thank me later*”; “*Potent combination of antiviral medicine for covid 19. 1. turmeric(a pinch)*.*2. panch tulasi(2-3 drops)*.*3. Indian borage leaf extract (2 spoons)*.*4. garlic paste or powder(1 spoon)*.*5. mint leaf extract (2 spoons)*.*6. honey (1 tablespoon)*.*7. salt(0*.*25 g)*.*cont*”

#### People Complaining about the side effects of taking herbs and vegetables

Not all was well with the users of herbs and vegetables as they took to twitter to complain of side effects. “*My internal organs reacting to me drinking my mom’s ginger; garlic; lemon; tumeric; and honey corona antivirus mixture; she picked up from facebook and whatsapp*”; “*To the whatsapp prof that told my mom ginger and garlic juice kills corona it doesn’t but it does kill the internal organs*”; “*When this COVID issue started; I trusted my ginger and garlic tea so much it became a daily routine to blend them fresh and steep in hot water. After a week of consistent consumption, I began to feel strange until I realize everything has its side effects*”; “*Can you ever take too much of ginger and vitamin c? cos now my tummy is turning me. Nitori corona oshi yii noni (It is because of stupid corona)*.”

#### Panic Purchase of certain herbs and vegetables

It is not a surprise that panic buy affected garlic, ginger, tumeric, lemon and other related products as people rush to stock the produce in large quantity. A respondent said, “*First time I’ve seen garlic and pancit canton packs sold out in grocery stores*”; “*All the garlic is gone. Not only can we not fight against covid-19 but vampires too*”; “*Am I seeing this correctly? There is no manuka honey left on any of the supermarket shelves. Apparently, it’s the magic cure/prevention of this virus all of sudden*”; “*Of the 10 things we tried to order today from our local provision store; 6 of them were out of stock. The out of stock items incl tea powder; dates; honey, chilli powder, tamarind etc*.”

#### Warning or correction on the fact that herbs and vegetables are not the cure for coronavirus

However, there are people that are monogamous enough to warn that consuming all these items are not enough to neither protect or cure people of COVID=19. Here are examples, *“Mom for the fifth time no u cannot cure corona with lemon tumeric ginger honey tea. “; “Lemon; garlic; ginger will not protect you against #covid19. There is in fact; no known cure or vaccine for covid-19. If you feel you have been exposed to covid-19 call 909 immediately”; “No; drinking hot lemon ginger garlic tea is not a cure for corona virus as told by a Chinese doctor on whatsapp. read along who’s myth busters:”; “There’s so many misinformation about covid-19 circulating in facebook. that ginger; garlic; salt; lemon; vitamin C; and many more; those are not a cure and does not have any scientific evidences. Vaccines are still at the process of clinic trials*.*”; “To the people here and on ig pedalling garlic; ginger and lemon concoctions as the cure for covid-19; please please please stop with this fake news. stopppp! It does not cure anything. Ko kin cure virus. Please add all other translations”; “Dear African parents; cutting onions in every room and putting garlic and ginger will not cure corona virus”; “experts say eating garlic does not prevent covid-19 -- and onions are no cure either”*

### Self-Medication Due to Prescription by Non-Medical Practitioners

This theme captures the conversations or expressions of people, who have got themselves involved in COVID-19 self-medication even without knowing their infection status. The thematic analysis will be around the following sub-themes: “Does an Influential COVID-19 Drug Recommendation Statement Matter?” and “Inquisitiveness of people on the usage of COVID-19 drugs Refuting the panacea”.“

#### Does an Influential COVID-19 Drug Recommendation Statement Matter?

President Donal Trump made public announcement on the fact that coronavirus can be treated with hydroxychloroquine was widely spread on the social media including tweeters. Here are examples, *“The president of the United States; Donald Trump; had announced on 19 march that chloroquine and hydroxychloroquine/plaquenil; used to treat malaria and arthritis; were approved by the FDA to be tested as a treatment for covid-19*.*”; “President Trump announced hydroxychloroquine and azithromycin (a broad spectrum antibiotics) on 03/21/20 for optional treatment of covid-19 infection. the efficacy of the hydroxychloroquine and broad spectrum antibiotic combination”; “Breaking news: Trump says he is swallowing the anti-malaria parasite drug; plaquenil (hydroxychloroquine); plus zinc every day; claiming unscientifically that it will be an effective preventive for”*. Based on this announcement people started taking the medicine without prescription and this led to self-poisoning as voiced out by a number of people. “*Another one here on issue of misusing #hydroxychloroquine. Two Nigerians overdose self-medicating with chloroquine after trump praised anti-malaria drug as possible covid-19 “; “Chloroquine did not poison anyone in Nigeria. Self-medication did. No doctor prescribed chloroquine neither did any doctor administer chloroquine to any patient infected with covid-19”;* “Cardiac toxicity cases have been reported in new aquitaine following self-medication of plaquenil (hydroxychloroquine) in the face of symptoms suggestive of covid-19; sometimes requiring hospitalization in intensive care.” We can see that the recommendation or endorsement of an influential person on the usage of a drug matters.

#### Inquisitiveness of people on the usage of COVID-19 drugs

Many people got information on certain drugs that can help in fight against the disease but are asking to verify the information that they received. Here is the range of questions on that. *“Does plaquenil or hydroxychloroquine users get covid 19?”; “Does hydroxychloroquine help covid patients. Has there been any trials? Is there any evidence?”; “But let me ask; what does hydroxychloroquine do to covid? “Can hydroxychloroquine protect patients with rheumatic diseases from covid-19? response to: ‘Does hydroxychloroquine prevent the transmission of covid-19?’”*.

#### Warning on self-medication of COVID-19 drugs

While some people have using COVId-19 drugs without medical guidance some people have been magnanimous enough to warn and advise people. Read some of their responses, *“#hcq should not be used for self-medication. it is to be given only under strict medical supervision*.*”, “Alert against the use of hydroxychloroquine in self-medication, cases of cardiac toxicity have been reported in new aquitaine in patients who have taken plaquenil without medical supervision in the face of symptoms suggesting a covid-19”. Certain medical practioner has and organisations have learnt their voices, “COVID-19: Self-medication of hydroxychloroquine is dangerous for patients with heart. Ulhas Pandurangi is a chief of division of cardiac electrophysiology & pacing at madras medical mission” “Don’t self-medicate by ingesting any of the following: hydroxychloroquine or chloroquine; bleach; hydrogen peroxide; excess colloidal silver; excess vitamin d; anything purported to be a covid-19 medication (there isn’t one*.*) this information comes from the Oregon poison center”*

### Conspiracy Theories on COVID-19 Theme

The thematic analysis around conspiracy theories focused on the following sub-themes (i) Conspiracy theory on the linkage between 5G technology and coronavirus (ii) Bill Gates and COViD-19 and (iii) The fabrication of coronavirus in a lab in China.

#### Linkage between 5G technology and coronavirus

We found people expressing evidence on people who believe that 5G is the cause of corona infection, “*On Jan 22nd a whole bloody doctor in Antwerp; claimed that 5G masts in Wuhan caused the corona virus. This conspiracy has spread like wildfire all over the net to the point where masts are getting burnt down in the UK FB; YouTube n Twitter keep taking down all these stories*” And some actually believe this theory to the extent of attacking telecommunication mast. Here is are statements on attacks of 5G towers or masts, “*The attacks stem from a conspiracy theory that 5G internet has caused the spread of the coronavirus*”; “*A conspiracy theory claiming that 5G internet has somehow caused the spread of the coronavirus has led to more than 70 cell phone towers in the United Kingdom to become targets of arson attacks; according to a report from Business Insider*”, “*20 Satellite masts were vandalised or set on fire across the UK this past weekend alone following false conspiracy theory claims that Chinese 5G caused Corona Virus*”.

#### Linking Bill Gates with the transmission of coronavirus

Many people believe the theory that Bill Gates is one of the brains behind COVID-19 infection and this is with intention making money from the sale of its vaccine. **“***Conspiracy theorists believe Bill Gates has something to do with this or at least their vaccinations will and they believe 5G caused COVID***”**. There was even a twitter poll whose result shows that a high number of people believe that Bill Gate is the cause of COVid-19. *“… results on a Twitter poll on this app where an alarming number of people believe that Bill Gates is out to sterilize the African continent and 5G caused COVID 19*”. There are some that demands investigation into the issue. “*Sir; I am from South Africa; I beg you to pls urgently investigate Bill Gates … everyone else involved in this Covid-19 conspiracy* …”; “…*Some Want to Investigate #CoronaConspiracy … #BillGatesFoundation*”.

#### Fabrication of coronavirus in a lab in China

It is all over the twitter space that coronavrisus was developed in a Chinese lab. *“The Chinese lab (conspiracy) theory is there a high-level bio lab in Wuhan”; “Would anyone really be surprised that a Chinese lab mishandled and mistakenly released the virus? and everyone knows china would not admit to this anyway. The Wuhan disease lab is the focus of suspicion and conspiracy theories about covid-19’s origins”*. This conspiracy theory was so strong that even there are sayings that president Trump and secretary of state bought into it. Here are some of the statements, *“Trump confident coronavirus may have originated in Chinese lab …”; “Trump says he has evidence coronavirus came from a Chinese lab”; “Trump says he has seen evidence that #coronavirus started in Wuhan virology laboratory”; “Pompeo says ‘enormous evidence’ coronavirus originated in Chinese lab”*.

### Fear and Anxiety Theme

This theme is all about the fear and anxiety that is associated with COVID-19. From Africa to Europe, there is fear in the hearts of people and their mouths are expressing them. Individuals, healthcare, workers are afraid of the virus.

#### Fear and anxiety everywhere

People are expressing general state of fear *“We are in a season where Fear; panic; anxiety; insecurity can be seen everywhere. The corona virus; suddenly appear and has become a big battle to human race. People are falling sick; and many people are at risk. Be encouraged and don’t be afraid. God will keep you SAFE”; “Why is everyone so afraid? Is it really because of covid? Or is it really because most of us are already living in a state of internal anxiety”*. The health care frontliners are afraid *“Today; the Covid positive patient I am caring for took a turn for the worse (ie. Comatose). The nurse I was on duty with last night has a cough and fever*.*I am afraid that any day I will be next. This is the fear and anxiety the frontliners likely share. I hope we beat this soon”*. People are afraid of going to hospital for non-coronavirus cases, “…*We have had many people afraid to attend hospital for NON COVID issues*”; “*By reaction to covid I mean this: People afraid to go to hospital; not been treated or screened. Private hospital services down due to lay off due to lack of patients. Depression; anxiety; suicides; alcoholism; drug abuse on the rise because of fear among other things*…” The children are afraid, “*My grandma was listening radio and they were mentioning that CHILDREN are afraid that THEY ARE GOING TO DIE BC OF THE CORONA VIRUS!*

*This is heartbreaking! Children should never ever fear for their lives! Please protect them and don’t let them read/watch the news all day*”. The elderly are afraid, “*Hearing now that the elderly are afraid to go out in public for fear of catching corona. Did Joe just lose his base?”*. The middle-aged people are afraid, “*The fear is real. It is not with the youth; elderly or those who have been working. But the middle aged*…” Young people are afraid, “*With the government focusing every effort to #COVID19KE young people are afraid of visiting health care systems to seek services as they fear contacting Covid 19*”

#### Fear and anxiety Mongering

Media is accused of instilling fear in the heart of people. *“Fear Mongering; the media overplayed the whole Corona virus and created fear and panic. Their end game was always to vaccinate you; but they needed your consent. We call that Problem-Reaction-Solution Paradigm. Agenda 2030 in summary”; “Fear-mongering techniques by the media is causing unnecessary panic wrt covid-19*.*” “COVID-19 anxiety is real and mutual fear-mongering; misinforming and denialism have become common social media responses to the pandemic”* People acknowledge the fact that fake news encourages fear mongering, “*At this rate; the covid virus will not be the one to kill people*…*people will be people’s downfall with fake news…”, “Fear is a bigger problem than the actual virus right now. Misinformation leads to fear; fear induces panic; and panic induces chaos. It needs to stop. The world is freaking out over this and for no good reason. At the moment the flu is more of a threat than corona virus*.*”*

#### Panic buying motivated by fear and anxiety

People are emptying the shelves buying more than what they need because they are afraid that other people will buy up everything and the fact that the government may put their communities, state and country may be put under locked down. People took to twitter to state this, “*Why the fuck are people panic buying toilet roll. Corona fear giving you the shits?*”; “*This whole panic buying of toilet roll is proof of how fear and panic can escalate out of control*.” Panic buying was limited to the developed countries of the world; the developing ones are also involved. Here are examples, “*Covid-19: Kano residents in panic buying over fear of shutdown*”; “*Panic buying escalates in Malaysia amid fear of Covid-19 lockdown*”.

#### Urban to rural movement

Even the rural areas are not left out in the impact of COVID-19 as people moving from urban to rural area of their countries. “*Wealthy people are rushing to move away from the big cities creating a real estate boom in many suburban areas; small towns and rural communities*”; “*People in Japan discussing COVID spread due to people leaving major metros for rural areas*”; ““

## Discussion

There is vast amount of information on various social media platforms such as twitter on COVID-19 pandemic and this ranges from fear and anxiety of people in contracting the disease and how they have resulted to measures that will either prevent them from getting infected or cure them of the infection. Studying COVID-19 from multifaceted perspectives is very import as the number of death from COVID-19 has overwhelmingly exceeded the toll of other disease outbreak in history(Baud et al., 2020; Weiss & Murdoch, 2020). It has kept the health system of most countries on their feet. In spite of plethora of studies on COVID-19 and its attendant consequences, there is less study on the perspectives of people on it. This study makes a unique contribution by performing thematic analysis of the opinion of people on social media about COVID-19. Four themes were identified from COVID-19 tweets: Herbs and Vegetables as COVID-19 Panacea, Self-Medication Due to Prescription by Non-Medical Practitioners on Social Media, Conspiracy Theories on COVID-19 and Fear and Anxiety attributed to the disease.

In this study, some of the tweets were misinforming people about the use of natural remedies like herbs and vegetables as a cure or prevention for COVID-19 disease. The spread of information on the social send people into panic buying of certain herbs and vegetables giving a sense of false hope and even combine them dangerously. This is not something new as people have suggested in the past that herbs such garlic, beetroot are the solution to HIV/AIDS (Baleta, 2006; Dzimiri, Dzimiri, & Batisai, 2019). It is noteworthy that a number of tweets show people warning against this belief and try dissuading people about the idea. Herbs and vegetables neither protect nor cure people of coronavirus (Khadka, Hashmi, & Usman, 2020; Phillips, 2020). However, compared to drugs, herbal products are not controlled for purity and potency. Rather, they are blended into foods and promptly accessible in the market without remedy (Abdel-Aziz, Aeron, & Khalil, 2016).

Also, many tweets about prescribing drugs by non-medical practitioners. Some of the tweets showed the high rise of curiosity on COVID-19 drug usage and also stern warning on the self-prescribing of drugs. However, the practice of self-medication is harmful. It could lead to some possible hazards like incorrect self-diagnosis, suspension in seeking timely medical advice,serious unfavourable reactions, risky drug interactions, risk of drug-dependence and abuse of drug use (Ruiz, 2010)

A good number of tweets conspired some unfound theories and fallacies associating technology and laboratories to the novel virus. That this COVID-19 is part of strategies conceived by some global elites like Bill Gate to make money from selling vaccine. Conspiracy is inclined to presume that notable public occasions are secretly arranged by influential and spiteful persons or organizations for certain gain (Douglas et al., 2019). There were many tweets relating 5G mobile network to breeding cononavirus. However, coronavirus is not transmitted through radio waves/mobile networks. There is also trend of tweets that this virus is only spread in countries with 5G mobile network which brought violence against telecommunication engineers and property. Jolley and Paterson (2020) also attested to the fact that this conspired of 5G and COVID-19 would breed anger, violence and paranoia. Several tweets also relating to the fabrication of coronavirus being a bio-weapon released deliberately by one of the laboratories at the Wuhan Institute of Virology in China. These kind of conspiracy tendencies have pushed people to discard established medicine extending to where once-cured diseases are making a rebound in some parts of the world. It also drove people to reject widely accepted scientific experiments (Douglas et al., 2019). Allington, Duffy, Wessely, Dhavan, and Rubin (2020) concluded that social media are one of the carriers of conspiracy belief which can only prevent health-protective behaviours.

A high number of people are afraid of COVID-19 pandemic. This is evidenced by the number of tweets on that. The effect of fear and anxiety on the said disease outbreak give rise to panic buying things like food and daily supplies, nose mask, face shield, hand sanitizers and potential medicines. Panic buying is categorized as survival psychology showing that behavioural changes can occur in people witnessing notable occurrences like catastrophes and disease outbreaks disrupting social lives or endanger health (Leach, 1994). Another subtheme from the many tweets show a large movement of people from urban to rural areas. Urban communities are at a greater danger of spread of diseases due to high population densities and extensive public transport network(WHO, 2020). The numerous tweets on pandemic fear and anxiety shows the mental health of people. Extreme cases of fear and anxiety are often triggered by mass tragedies like infectious disease outbreak which causes disruptions to the behaviour and psychological well-being of many in the population (Balaratnasingam & Janca, 2006)

## Conclusion

This study capitalises on social media, a discursive space in which individuals with or without experience of covid-19 shared information and their perspectives of COVID-19. In alignment with our study objective, we have been able to identify and analyse different themes (Herbs and Vegetables; Self Medication Due to Prescription by Non-Medical Practitioners; Conspiracy theory; Fear and Anxiety) about the pandemic. Social media has become a wider part of people’s lives and has large impact on decisions in people’s lives, it is important to research on its influence in people and health during this pandemic.

## Data Availability

Yes it is available

https://twitter.com/home

